# The addiction neurocircuitry during Resting-State Functional Connectivity fMRI in people with Cannabis User Disorder who tried to cut down or quit

**DOI:** 10.1101/2025.03.31.25324487

**Authors:** Hannah Thomson, Izelle Labuschagne, Arush Honnedevasthana Arun, Eugene McTavish, Hannah Sehl, Adam Clemente, Emillie Beyer, Marianna Quinones-Valera, Peter Rendell, Gill Terrett, Lisa-Marie Greenwood, Govinda Poudel, Victoria Manning, Chao Suo, Valentina Lorenzetti

## Abstract

Cannabis use disorder (CUD) affects ∼22M people globally and is characterised by difficulties in cutting down and quitting use, but the underlying neurobiology remains unclear. We examined resting-state functional connectivity (rsFC) between regions-of-interest (ROIs) of the addiction neurocircuitry and the rest of the brain in 65 individuals with moderate-to-severe CUD who reported attempts to cut down or quit, compared to 42 controls, and explored the association between rsFC and cannabis exposure and related problems, to elucidate potential drivers of rsFC alterations. The CUD group showed greater rsFC than controls between ROIs implicated in reward processing and habitual substance use (i.e., nucleus accumbens, putamen, pallidum) and occipito/parietal areas implicated in salience processing and disinhibition. Putamen-occipital rsFC correlated with levels of problematic cannabis use and depression symptoms. CUD appears to show neuroadaptations of the addiction neurocircuitry, previously demonstrated in other substance use disorders.

## INTRODUCTION

Cannabis Use Disorder (CUD) is experienced by ∼22 million individuals globally (Alcohol & Drug Use, 2018), and can be characterised by an inability to voluntarily cease or reduce consumption of cannabis despite experiencing physical or psychological harms (American Psychiatric Association [APA], 2013), e.g., risk-taking behaviours, including driving under the influence (Australian Institute of Health and Welfare [AIHW], 2020). CUD is associated with a greater prevalence of mental health disorders such as depression, anxiety, and psychosis (Jefsen et al., 2023; Lawn, Mokrysz, et al., 2022) and reduced performance on cognitive tasks (Lawn, Fernandez-Vinson, et al., 2022; Lorenzetti et al., 2020). Psychosocial problems associated with CUD have been partly ascribed to neurobiological changes within the addiction neurocircuitry (Koob & Volkow, 2016). Of relevance, recent functional neuroimaging methodologies have measured the functional integrity of the addiction neurocircuitry *in vivo*, minimising confounding effects of task-related cognitive processes associated with task performance, e.g., greater cognitive effort and the involvement of task-specific cognitive processes (Smitha et al., 2017). One such methodology is resting-state functional connectivity (rsFC). rsFC measures correlations between the spontaneous fluctuations of the blood oxygen level-dependent (BOLD) signal of two or more spatially distinct brain regions while people are at rest and without performing cognitively demanding tasks (Greicius et al., 2009; Smitha et al., 2017; van de Ven et al., 2004). Investigating rsFC in CUD can be valuable to offer novel insights into how the integrity of the addiction neurocircuitry is affected during rest and when the system is not challenged by meeting the demands of specific cognitive tasks.

Mounting evidence from functional Magnetic Resonance Imaging (fMRI) studies demonstrates that people who regularly use cannabis show different brain function during rest (i.e., rsFC) – within brain pathways of the addiction neurocircuitry (Koob & Volkow, 2016). Compared to controls, they show greater rsFC between frontal regions and: other frontal regions (e.g., anterior cingulate cortex [ACC], prefrontal and orbitofrontal cortices [PFC & OFC]), temporal regions (e.g., hippocampus, amygdala), and striatal regions (e.g., nucleus accumbens [NAc], putamen, pallidum, caudate; Thomson et al., 2022). Researchers have proposed that identified changes may involve circuitry important for cognitive functions commonly altered in CUD e.g., greater salience of cannabis rewards, lower sensitivity to rewards other than cannabis, disinhibition, stress (Thomson et al., 2022). There is also preliminary evidence that rsFC changes in cannabis users correlated with cannabis exposure metrics (i.e., age of use onset, duration; Thomson et al., 2022); however, the significance and direction of the findings have been inconsistent and require further investigation.

The evidence on rsFC changes in cannabis users is limited by methodological issues. First, only two fMRI rsFC studies, to the best of our knowledge, have measured if their participants endorsed DSM-5 criteria for CUD (APA, 2013; Aloi et al., 2021; Kroon et al., 2024). In both studies, rsFC significantly differed between CUD and control groups and correlated with metrics of cannabis use (Structured Clinical Interview of DSM-5 – research version [SCID-5-RV] scores, Cannabis Use Disorders Identification Test – Revised [CUDIT-R] scores, and cannabis grams per week), supporting the importance of considering both CUD status and levels of cannabis use. However, the majority of literature in this field does *not* account for CUD status and has not formally selected participants with more severe forms of CUD with failed attempts to cut down or quit; hence, the extant literature’s relevance for people with CUD remains unclear (Thomson et al., 2022). This issue is important as a substantial number of individuals who use cannabis report failed attempts to quit (Hughes et al., 2016), and the underlying neurobiology remains unexamined. Second, the impact of confounding variables that can affect brain function independently or in interaction with CUD, has been inconsistently examined, including key sociodemographic data (e.g., age and sex), alcohol and nicotine consumption, and mental health symptoms (e.g., depression; Thomson et al., 2022). Third, cannabis exposure and related problems have seldom been measured in relation to rsFC. Therefore, it remains unclear if these factors play a role in driving altered rsFC reported in cannabis users (e.g., severity of CUD, dosage of recent cannabis use, urine levels of 11-nor-9-carboxy-Δ□-tetrahydrocannabinol: creatinine [THC-COOH: creatinine ng/ml], and duration of cannabis use and of abstinence). Finally, about half of the extant literature consists of small sample sizes (i.e., *n* < 25; Thomson et al., 2022), and may be statistically underpowered to detect subtle rsFC alterations.

We aimed to overcome these limitations by examining rsFC in a sample of 107 participants (35 females and 72 males) aged 18-to-56. The primary aim was to examine rsFC in 65 individuals with moderate-to-severe CUD who had recently tried to cut down/cease cannabis use, compared to 42 controls, accounting for age, sex, and standard drinks in the previous 30 days. It was hypothesised that compared to controls, the CUD group would show different rsFC between regions-of-interest (ROIs) and the rest of the brain, utilising a seed-to-whole brain analysis. ROIs were concurrently integral to the addiction neurocircuitry (Koob & Volkow, 2016) and with known rsFC alterations in cannabis users (Thomson et al., 2022). Said ROIs were also dense in cannabinoid receptors type 1 (CB_1_R), to which delta-9-tetrahydrocannabinol (THC), the main psychoactive compound of cannabis binds to (Glass et al., 1997; Zou & Kumar, 2018). The selected ROIs were in the striatum (i.e., NAc, putamen, caudate), basal ganglia (pallidum), medial temporal regions (i.e., hippocampus, amygdala), and the ACC. By targeting these specific regions within the addiction neurocircuitry and investigating their connectivity with the rest of the brain, the findings aim to enhance our understanding of the neurobiological underpinnings of CUD.

The secondary aim was to explore how significantly altered rsFC in the cannabis group correlated with metrics of cannabis exposure and related problems, including SCID-5-RV scores, CUDIT-R scores, grams/past 30 days, urine levels of THC-COOH: creatinine ng/ml, hours since last use, age of onset, duration of regular use, depression symptom scores, nicotine dependence (Aloi et al., 2021; Blanco-Hinojo et al., 2017; Filbey et al., 2014; Kroon et al., 2024; Wetherill et al., 2015). By examining these relationships, the study aims to uncover potential drivers of rsFC changes, offering deeper insights into the neurobiological mechanisms of CUD.

## METHODS

### Study Design

This cross-sectional, case-control study was nested within a larger, pre-registered project (ISRCTN ID: 76056942) and was approved by the Australian Catholic University Human Research and Ethics Committee (HREC:2019-71H).

### Participants

#### Recruitment

All participants were recruited from the Greater Melbourne Metropolitan area via community-based flyers and online advertisements, which directed them to an online screening survey. Participants were further screened via phone call to confirm eligibility for the study.

#### Key inclusion and exclusion criteria

Participants aged 18-55^a^ were eligible if they *either* endorsed moderate-to-severe CUD (APA, 2013), with use of cannabis on a daily or almost daily basis for ≥12 months, and with at least one attempt to quit or reduce cannabis use within the past 24 months; *or* were controls who *did not* endorse the use of cannabis at any stage in the 12 months prior to testing, or >50 lifetime uses of cannabis. A complete and detailed overview of the inclusion and exclusion criteria is displayed in the Supplementary Information.

### Procedure

Participants provided written informed consent before completing a ∼4-6-hour session that included demographic surveys, fMRI scanning, behavioural and cognitive assessments, and urine sample collection. The sessions were conducted by experienced and specially trained researchers and students at the Monash Biomedical Imaging facility in Clayton, Victoria. Please see Supplementary Information for detailed participant procedures.

### Measures

All measures were described in detail in the study pre-registration (https://doi.org/10.1186/ISRCTN76056942). Participants’ age, sex, and years of education were measured using a standard demographic proforma. Handedness was ascertained using the Edinburgh Handedness Inventory – Short Form (EHI-SF; Veale, 2014). The Wechsler Abbreviated Scale of Intelligence – second edition (WASI-II; Wechsler, 2011) was used to estimate participants’ FSIQ, derived from administering the Vocabulary and Matrix Reasoning subtests.

Presence of at least moderate CUD was established using the SCID-5-RV (First, 2015). Participants were required to respond ‘yes’ or ‘no’ to 11 items to determine the number of CUD symptoms endorsed. A score of 4-5 indicated moderate CUD, and a score of 6+ indicated severe CUD. Semi-structured interviews were administered to measure past-30-day-use of any substances (Timeline Follow Back [TLFB]; Sobell et al., 1986); and lifetime cannabis use (Lorenzetti et al., 2015; Solowij et al., 2002; Yucel et al., 2008). From these, days and dosage of cannabis, alcohol, and nicotine (i.e., grams, alcohol, and number of cigarettes; TLFB) were extracted, as well as age of regular cannabis use onset and duration of regular use, factoring in self-reported use breaks; regular use was defined as at least three times per week. Substance use and related problems were quantified for cannabis, alcohol, and nicotine using the CUDIT-R (Adamson et al., 2010), the Alcohol Use Disorders Identification Test (AUDIT; Babor et al., 2018) and the Fagerström Test for Nicotine Dependence, respectively (FTND; Fagerström et al., 2012). Urine levels of THC metabolites were also analysed i.e., THC-COOH: creatinine ng/ml.

Several scales quantified mental health symptom scores. Depression was measured via the Beck Depression Inventory – second edition (BDI-II; Beck et al., 1996), state anxiety was assessed with the State-Trait Anxiety Index – Y Form, ‘state’ sub-scale (STAI-Y; Spielberger et al., 1983), and sub-clinical positive psychotic symptoms, negative psychotic symptoms and depressive symptoms were gauged with the Community Assessment of Psychic Experiences (CAPE; Stefanis et al., 2002). Further, stress levels were measured using the Perceived Stress Scale (PSS) – 10 items (Cohen et al., 1983), while the COVID Stress Scale estimated COVID-specific stress (Taylor et al., 2020).

### fMRI Acquisition and Analysis

MRI data acquisition, pre-processing, quality control and analysis are detailed in the Supplementary Information. Key acquisition parameters were as follows: TE = 2.07ms, TR = 2300ms, flip angle = 9, field of view 256 x 256mm, 192 sagittal slices without gap, yielding a 1 x 1 x 1mm resolution, with a total acquisition time of 5 minutes. Resting-state fMRI scans (189 volumes) were acquired over 8 minutes using T2* weighted Echo Planar Imaging (EPI) with TE = 30ms, TR = 2500ms, flip angle = 90, field of view = 192mm, 44 slices without gap, matrix = 64, and a voxel size of 3mm^3^. Standard pre-processing pipeline were conducted using CONN toolbox 20.b (Whitfield-Gabrieli & Nieto-Castanon, 2012) on a cloud-based computational platform (MASSIVE; Goscinski, 2014). *A priori* brain ROIs were selected as seeds: NAc, putamen, pallidum, caudate, hippocampus, amygdala and the ACC. Figure 1 overviews the seeds examined. Functional connectivity maps for each seed were generated for each individual and fed into the next statistical analysis.

**Figure 1.**
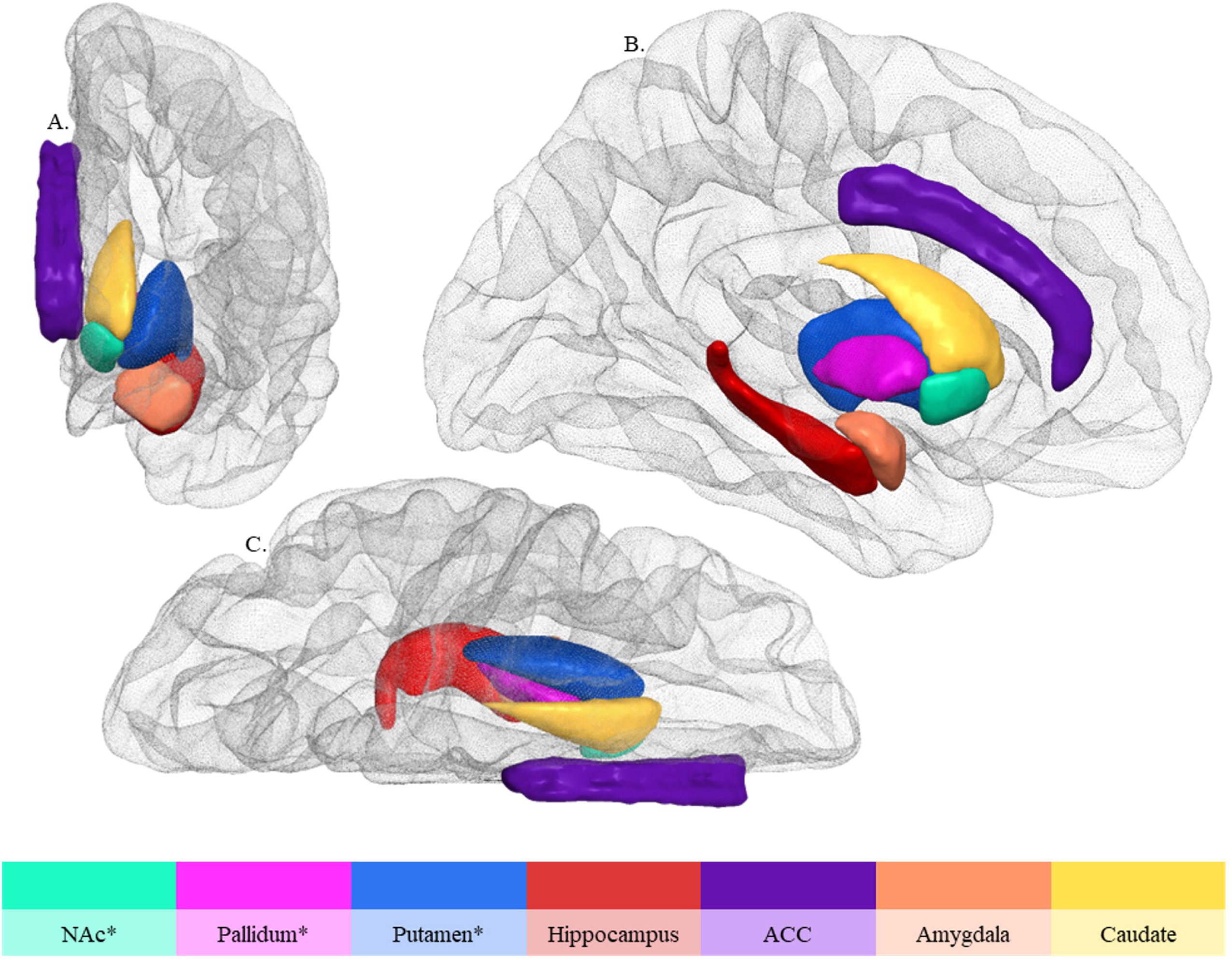
Overview of examined seeds, shown here in the *left* hemisphere only. **\***Ultimately found to significantly functionally connect to other brain voxels A., anterior view; ACC, anterior cingulate cortex; B., sagittal view; C., superior view; NAc, nucleus accumbens

### Statistical Analyses

#### Sample Characteristics

Groups were compared using Chi-squares for categorical variables (i.e., sex, handedness), t-tests for normally distributed scalar data (i.e., FSIQ, perceived stress), and Mann-Whitney U tests for other non-normally distributed scalar data.

#### Seed-to-Whole brain resting-state Functional Connectivity analysis

Statistical analysis was conducted using the same CONN toolbox to examine group differences for seed-to-whole brain connectivity, adjusting for age, sex, and past 30-days standard drinks. Two corrections for multiple comparisons were applied before significant ROIs were reported. Specially, a false discovery rate (FDR) correction was applied to control for voxel-wise multiple comparison errors, and a Bonferroni correction was applied for independent seed selections. Please refer to Supplementary Information for details.

#### Brain-Behaviour Correlations

The rsFC values within each significant ROI were extracted at the individual level. Spearman’s correlations examined the relationship between post-hoc ROI values, and cannabis exposure and related problems: number of DSM-5 CUD symptoms, CUDIT-R scores, cannabis grams/past 30 days, THC-COOH: creatinine ng/ml in urine, hours since last cannabis use, age of onset, duration of regular cannabis use, depression symptoms, and FTND. Correlations’ p-values were Bonferroni-corrected to *p* < .001 after dividing *p* < .05 by 45 correlations run. All descriptive statistics and brain-behaviour correlations were run via SPSS version 28.

## RESULTS

### Sample Characteristics

Sample characteristics are outlined in Table 1. The sample included 107 participants (65 with moderate-to-severe CUD, 42 controls) aged 18–56 years (mean = 27, 35 females and 72 males). Groups were matched by sex and age. The CUD and control groups did not significantly differ for years of education, FSIQ estimates, and handedness, as well as for several mental health symptom scores: state anxiety, perceived stress, and COVID stress.

**Table 1.**
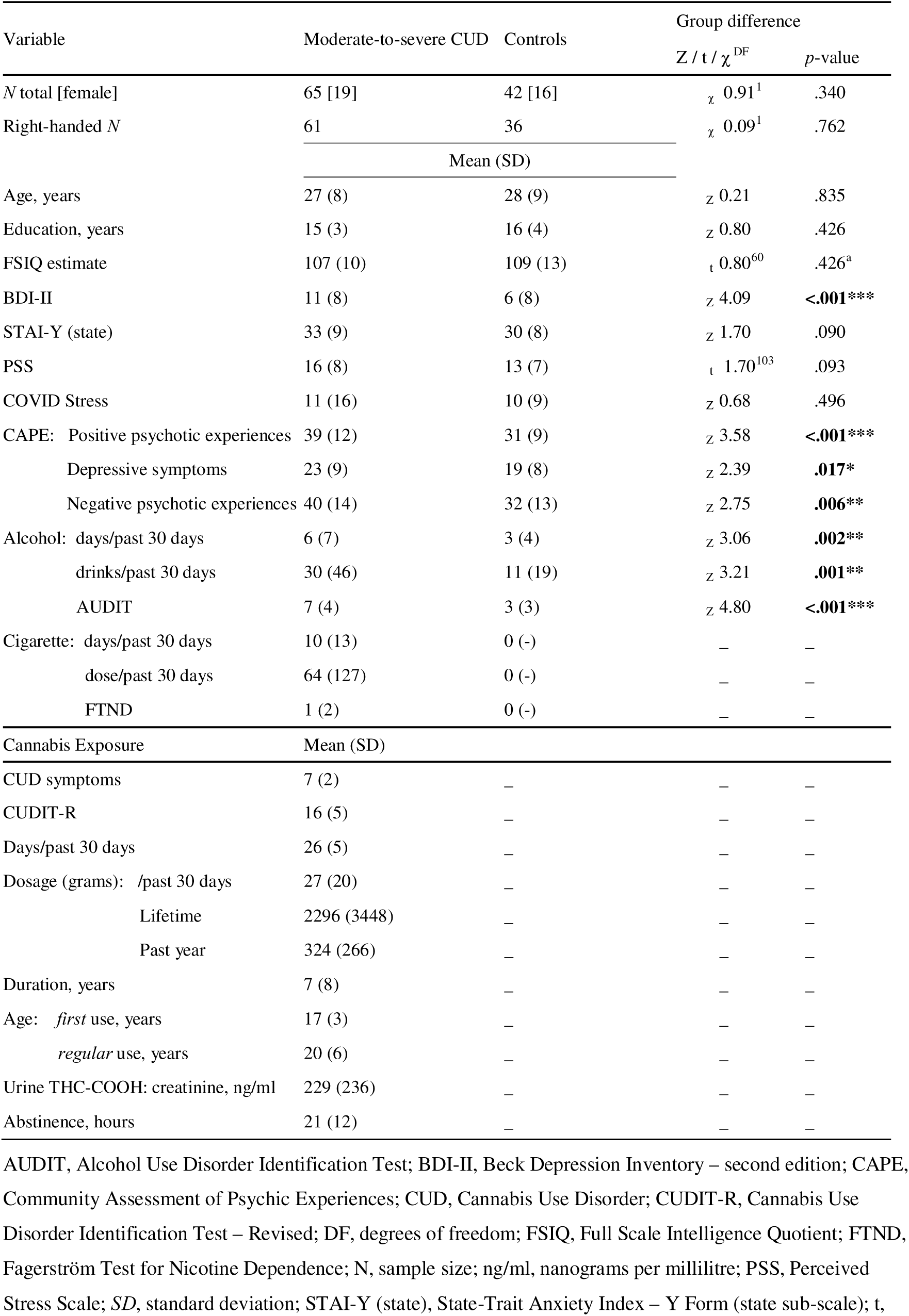
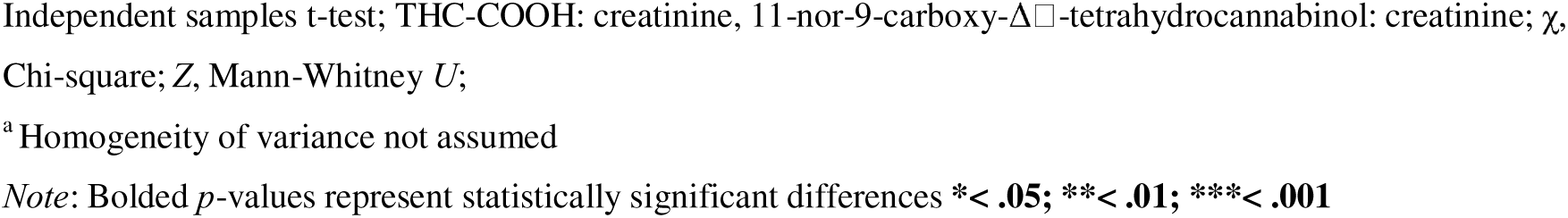
Sample characteristics and group differences for sociodemographic data, FSIQ estimate, substance use and related problems, and mental health symptom scores.

| Variable | Moderate-to-severe CUD | Controls | Group difference |  |
| --- | --- | --- | --- | --- |
| | | | Z / t / $\chi^2$<sup>DF</sup> | p-value |
| N total [female] | 65 [19] | 42 [16] | $\chi^2$ 0.91<sup>1</sup> | .340 |
| Right-handed N | 61 | 36 | $\chi^2$ 0.09<sup>1</sup> | .762 |
|  | Mean (SD) |  |  |  |
| Age, years | 27 (8) | 28 (9) | Z 0.21 | .835 |
| Education, years | 15 (3) | 16 (4) | Z 0.80 | .426 |
| FSIQ estimate | 107 (10) | 109 (13) | t 0.80<sup>60</sup> | .426<sup>a</sup> |
| BDI-II | 11 (8) | 6 (8) | Z 4.09 | <.001*** |
| STAI-Y (state) | 33 (9) | 30 (8) | Z 1.70 | .090 |
| PSS | 16 (8) | 13 (7) | t 1.70<sup>103</sup> | .093 |
| COVID Stress | 11 (16) | 10 (9) | Z 0.68 | .496 |
| CAPE: Positive psychotic experiences | 39 (12) | 31 (9) | Z 3.58 | <.001*** |
| Depressive symptoms | 23 (9) | 19 (8) | Z 2.39 | .017* |
| Negative psychotic experiences | 40 (14) | 32 (13) | Z 2.75 | .006** |
| Alcohol: days/past 30 days | 6 (7) | 3 (4) | Z 3.06 | .002** |
| drinks/past 30 days | 30 (46) | 11 (19) | Z 3.21 | .001** |
| AUDIT | 7 (4) | 3 (3) | Z 4.80 | <.001*** |
| Cigarette: days/past 30 days | 10 (13) | 0 (-) | – | – |
| dose/past 30 days | 64 (127) | 0 (-) | – | – |
| FTND | 1 (2) | 0 (-) | – | – |
| Cannabis Exposure | Mean (SD) |  |  |  |
| CUD symptoms | 7 (2) | – | – | – |
| CUDIT-R | 16 (5) | – | – | – |
| Days/past 30 days | 26 (5) | – | – | – |
| Dosage (grams): /past 30 days | 27 (20) | – | – | – |
| Lifetime | 2296 (3448) | – | – | – |
| Past year | 324 (266) | – | – | – |
| Duration, years | 7 (8) | – | – | – |
| Age: first use, years | 17 (3) | – | – | – |
| regular use, years | 20 (6) | – | – | – |
| Urine THC-COOH: creatinine, ng/ml | 229 (236) | – | – | – |
| Abstinence, hours | 21 (12) | – | – | – |

AUDIT, Alcohol Use Disorder Identification Test; BDI-II, Beck Depression Inventory – second edition; CAPE, Community Assessment of Psychic Experiences; CUD, Cannabis Use Disorder; CUDIT-R, Cannabis Use Disorder Identification Test – Revised; DF, degrees of freedom; FSIQ, Full Scale Intelligence Quotient; FTND, Fagerström Test for Nicotine Dependence; N, sample size; ng/ml, nanograms per millilitre; PSS, Perceived Stress Scale; SD, standard deviation; STAI-Y (state), State-Trait Anxiety Index – Y Form (state sub-scale); t,
Independent samples t-test; THC-COOH: creatinine, 11-nor-9-carboxy- $\Delta^9$ -tetrahydrocannabinol: creatinine; $\chi^2$ , Chi-square; Z, Mann-Whitney *U*;
<sup>a</sup> Homogeneity of variance not assumed
*Note:* Bolded *p*-values represent statistically significant differences \***< .05**; \*\***< .01**; \*\*\***< .001**

The CUD group had significantly higher scores than controls for depression symptoms and all CAPE subscales, as well as all alcohol use metrics: alcohol use days/past 30 days, standard drinks/past 30 days, and AUDIT scores.

There were 28 of 65 people with CUD who endorsed nicotine use over the past 30-days; they scored a mean of 2 on the FTND, indicating ‘very low’ nicotine dependence overall. No controls reported past 30-day nicotine use.

### Levels of Cannabis Exposure and Related Problems

All people in the CUD group met the criteria for moderate-to-severe CUD; on average, participants endorsed severe CUD, corroborated by DSM-5 CUD symptoms. Participants used cannabis almost daily over the past 30 days and about a gram per day.

All people with CUD reported having attempted to cut down or reduce their cannabis use at least once over the past 2 years and used cannabis at least 4 days per week for a minimum of 12 months. The mean age of *first* cannabis use was 17, and of *regular* use was 20.

The CUD group abstained from cannabis for a mean of 21 hours before testing. The presence of THC metabolites in urine confirmed cannabis use in the CUD group, accounting for variable hydration status.

### Group Differences in Resting-State Functional Connectivity

As shown in Table 2 and Figure 2, the CUD group had higher rsFC than controls between fronto-striatal, occipito-parieto-striatal and occipito-basal ganglia region pairings, accounting for age, sex, and past 30-day standard drinks, with FDR and Benjamini-Hochberg (1995) corrections applied. Specific region pairings showing greater rsFC were: NAc-frontal pole/SFG, putamen-occipito/parietal; and pallidum-occipital/intracalcarine cortex.

**Figure 2.**
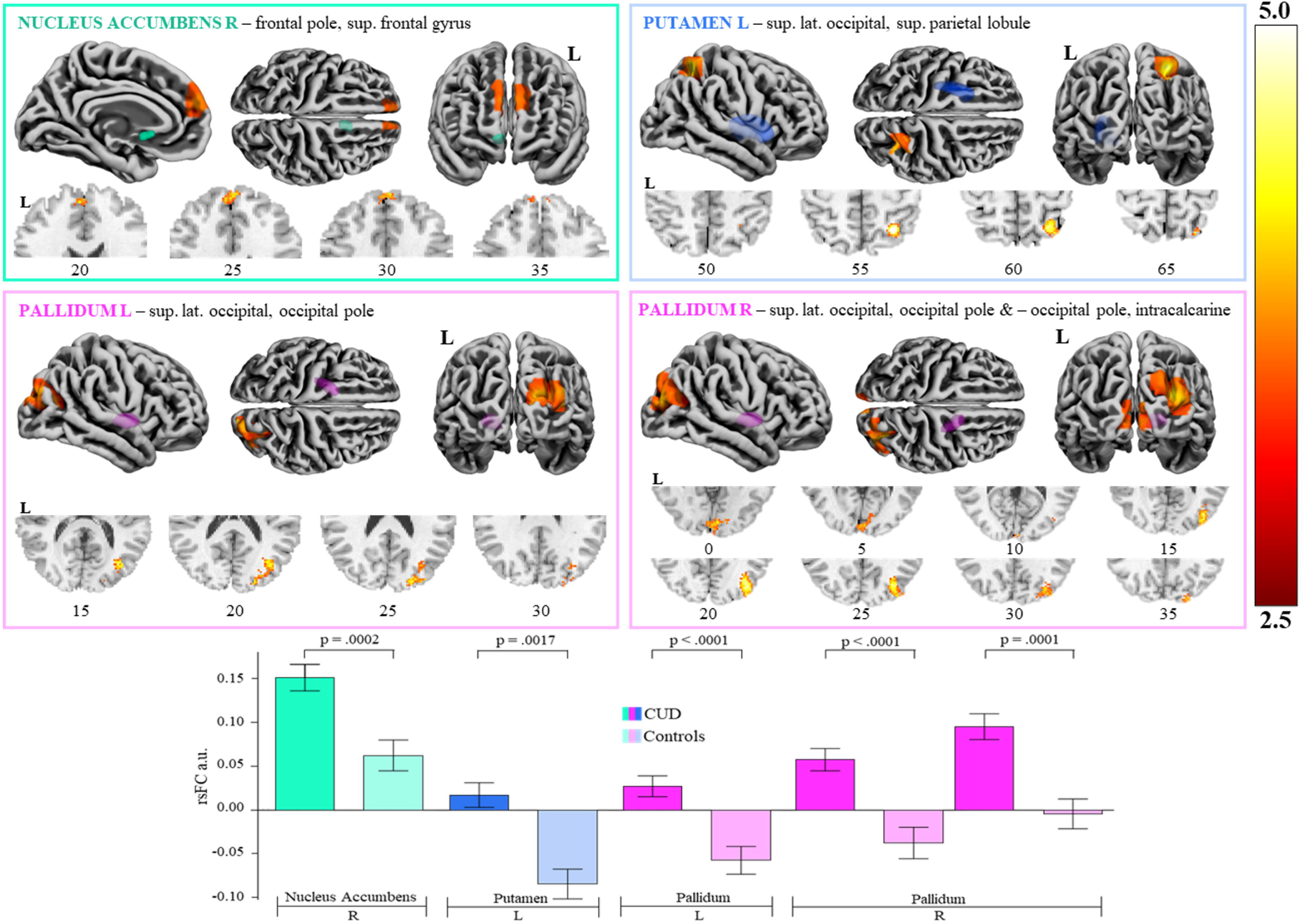
Significantly greater resting-state functional connectivity in people with a Cannabis Use Disorder compared to controls, between ROI seeds & clusters. a.u., arbitrary units; CUD, cannabis use disorder; L, left; lat., lateral; rsFC, resting-state functional connectivity; ROI, region of interest; sup., superior

**Table 2.** Overview of significantly greater resting-state functional connectivity in people with CUD compared to controls, between ROI seeds and their respective clusters.

| Seed | Cluster (*specific regions*) | Peak (x, y, z) | K | Size p-FDR | B-H adjusted $\alpha$ |
| --- | --- | --- | --- | --- | --- |
| NAc, *right* | frontal pole *bilateral*, SFG *left* | 6, 54, 22 | 190 | **.0002\*\*\*** | .0133 |
| putamen, *left* | superior lateral occipital, superior parietal lobule *right* | 30, -56, 56 | 178 | **.0017\*\*** | .0167 |
| pallidum, *left* | superior lateral occipital, occipital pole *right* | 36, -74, 18 | 317 | **<.0001\*\*\*** | .0067 |
| *right* | superior lateral occipital, occipital pole *right* | 34, -76, 24 | 501 | **<.0001\*\*\*** | .0033 |
|  | occipital pole, intracalcarine *bilateral* | 4, -90, 2 | 229 | **.0001\*\*\*** | .0100 |

B-H, Benjamini-Hochberg; FDR, false discovery rate; K, number of voxels; NAc, nucleus accumbens; SFG, superior frontal gyrus
*Note:* Bolded *p*-values represent statistically significant pairings \*\*< .01; \*\*\*< .001

### Brain-Behaviour Correlations

Significant correlations were observed between greater left pallidum-occipital cortex rsFC and lower CUDIT-R scores (*p* =. 0007) and lower depression symptom scores (i.e., BDI-II, *p* = .0004), following the application of a Bonferroni correction (Figure 3). No other correlations reached statistical significance (see Supplementary Table 1).

**Figure 3.**
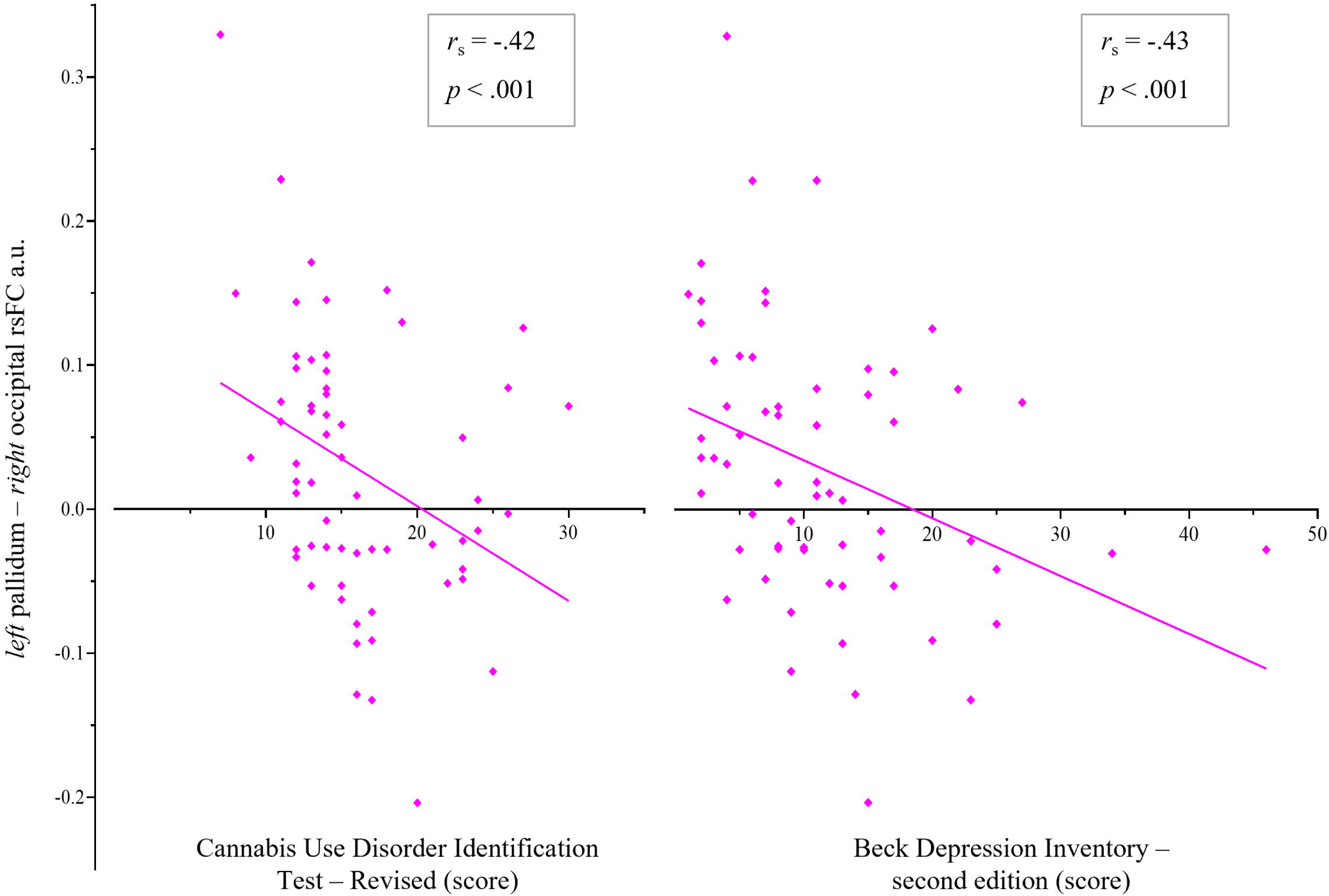
Spearman’s rho (r_s_) correlations within the CUD group, between rsFC pairs showing group differences (rsFC beta values) significantly correlated with metrics of cannabis exposure and related problems and depression scores, using Bonferroni adjusted alpha threshold of p < .001 (.05 ÷ 45)

## DISCUSSION

To our knowledge, this is the first rsFC fMRI study to examine individuals with moderate-to-severe CUD who have attempted to cut down or cease cannabis use, while controlling for age, sex, and recent alcohol use. Consistent with prominent neuroscientific theories of addiction, CUD was shown to be associated with altered rsFC across striatal and basal ganglia sub-cortical regions, and frontal, occipital, and parietal cortical regions compared to controls. Identified regions have been implicated in reward processing, disinhibition, and habitual/compulsive substance use (i.e., NAc-frontal pole/SFG, putamen-occipito/parietal cortex, pallidum-occipital/intracalcarine cortex); some of which correlated with severity of CUD scores and depression symptom scores.

The current study consistently observed greater rsFC in CUD compared to controls, particularly within fronto-striatal and occipito-parieto-striatal networks. The location and direction of the findings are in line with previous work reporting greater rsFC in overlapping networks in people who regularly use cannabis compared to controls (i.e., greater NAc-ACC cluster rsFC; Zhou et al., 2018). The direction of rsFC change may fluctuate as a function of the intoxication cycle. Indeed, greater rsFC has been consistently reported when participants are non-intoxicated (Ertl et al., 2023), while lower rsFC has been consistently reported during acute cannabis intoxication (Wall et al., 2019). Therefore, chronic intoxication with THC over time may lead to greater connectivity of mesocorticolimbic regions (e.g., between frontal regions and the motor cortex, insula, and temporo-parietal regions; Ertl et al., 2023).

We found greater putamen-occipito/parietal and pallidum-occipital/intracalcarine rsFC in CUD compared to controls. This pattern of rsFC, may underlie habitual cannabis use and altered salience processing observed in CUD (Everitt & Robbins, 2005). Indeed, both the putamen and pallidum have been implicated in cognitive processes reportedly altered in regular cannabis users, including habit formation/automated behaviour (Everitt & Robbins, 2005; Grahn et al., 2008; Yin & Knowlton, 2006) and habitual substance use (Everitt & Robbins, 2005).

The observed correlation between greater pallidum-occipital cortex rsFC and lower CUDIT-R scores contributes to the literature which suggests that rsFC changes may reflect neural adaptations linked to cannabis use severity (Aloi et al., 2021; Kroon et al., 2024). These findings align with previous evidence implicating the pallidum and putamen in habit formation and compulsive substance use (Everitt & Robbins, 2005). This notion is partially supported by the association identified herein between pallidum-occipital cortex rsFC and CUDIT-R scores. Similarly, greater activation in striatal and basal ganglia regions has emerged in cannabis users with greater severity of cannabis-related problems, frequency of use and/or earlier age of onset across task-based ‘cue-reactivity’ fMRI research (Sehl et al., 2021). In contrast, task-based fMRI studies have also detected decreased activity in the putamen and in occipital regions, in long-term cannabis users (Blest-Hopley et al., 2018). Occipital (and some parietal) regions support attentional processing/direction toward salient information such as ‘uniquely coloured lines’, or specific-coloured circles, presented in the participant’s visual field (Beffara et al., 2022; Gilbert & Li, 2013; Kim et al., 2021), which could reflect neuroadaptations in attention/thoughts towards cannabis/salience pathways that persist during rest.

The notion that established rsFC alterations in a cannabis-using group may be driven in part by the presence/severity of CUD (Lorenzetti et al., 2016), is supported by the negative association identified between pallidum-occipital cortex rsFC and CUDIT-R scores. Alterations in such neural structures are thought to underlie emotional processing, habit formation, and reward-based learning. Additionally, this correlation was largely aligned with correlations reported by Aloi et al. (2021), who demonstrated a negative association between CUDIT-R scores and amygdala-occipital (cuneus) rsFC. Negative correlations between CUDIT-R scores and rsFC have also been reported for the following pairings; crus-brain stem and crus-cerebellar lobule (Sweigert et al., 2020), and amygdala-paracentral/supplementary motor area and amygdala-temporal pole (Aloi et al., 2021). Additionally, negative associations established between pallidum-occipital cortex rsFC and depression scores were also somewhat in keeping with previous research (Shollenbarger et al., 2019; Subramaniam et al., 2018). Depression scores were previously shown to positively correlate with rsFC between frontal regions (Shollenbarger et al., 2019) and between frontal-parietal regions (Subramaniam et al., 2018). These findings highlight the importance of controlling for mood-related symptomology when examining rsFC in cannabis-using populations. It also suggests that people with comorbidities (i.e., mood disorder), may experience additional neuroadaptations and hence may be an especially vulnerable subgroup of people with CUD.

We reported greater fronto-striatal (NAc-frontal pole/SFG) rsFC in people with CUD, which may reflect increased engagement of salience pathways sensitive to THC exposure (Berridge & Robinson, 2016). Indeed, THC increases dopamine synthesis within the NAc (Bossong et al., 2009; Pierce & Kumaresan, 2006), which might subsequently affect the *function* of NAc, and possibly of interconnected frontal pathways implicated in salience processing. It is acknowledged that we cannot ascertain which region drives the connection or if both/all regions activate in unison, from the identified rsFC alterations. Nevertheless, increased fronto-striatal rsFC may be a key contributor to cannabinoid reinforcement (Lupica et al., 2004; Tanda & Goldberg, 2003). Indeed, neural projections from the NAc to the PFC may mediate the experience of ‘wanting’ cannabis and urges to use cannabis (Berridge & Robinson, 2016). In line with this notion, the NAc plays a key role in assigning value to stimuli (Knutson & Gibbs, 2007), whilst frontal regions have been linked to a loss of control over substance use (George & Koob, 2010). Therefore, fronto-striatal regions may be more prominently implicated in more severe forms of CUD, where a loss of control over cannabis use can be a key feature.

Overall, our finding of greater fronto-striatal connectivity in people with more severe forms of CUD is in keeping with rsFC fMRI literature in cannabis users more broadly (for a review, see Thomson et al. [2022]) and in studies of other substance use disorders (SUDs). Specifically, greater frontostriatal rsFC was observed in current, ‘chronic’ cannabis users (Blanco-Hinojo et al., 2017), in cannabis-dependent participants who abstained from cannabis for 28 days (Zhou et al., 2018; Zimmermann et al., 2018), and in users of different substances: cocaine (Hu et al., 2015; Zhang & Li, 2018), nicotine (Hong et al., 2009; Hong et al., 2010), opioids (McConnell et al., 2020; Wang et al., 2021), stimulants (Wang et al., 2019), and polysubstance (Motzkin et al., 2014). Taken together, the findings suggest that fronto-striatal rsFC alterations may underpin CUD, although the time course of the changes is unclear as they may predate or follow CUD (or both); and may persist following abstinence from cannabis.

To the authors’ knowledge at the time of writing, this is the first fMRI study to examine rsFC in current cannabis users with moderate-to-severe CUD who report failed attempts to cut down or quit their consumption. Findings herein lend support for the first time, that the neurocircuitry of CUD overlaps with the addiction neurocircuitry implicated in reward/salience processing and compulsive use, as postulated by prominent neuroscientific theories (Berridge & Robinson, 2016; Koob & Volkow, 2010; Volkow et al., 2016), which have largely been based on substances other than cannabis (e.g., cocaine, alcohol, opioids; Koob & Volkow, 2010; Volkow et al., 2016). Additionally, up until now, established neuroscientific theories of addiction have been largely validated in outdated diagnostic systems (e.g., DSM-IV), which do not reflect the current diagnostic system relying on the DSM-5 (Livne et al., 2021). Furthermore, we have specifically examined cannabis users who endorse an inability to reduce or cease their use, extending our findings to a large subset of individuals with CUD (Hughes et al., 2016). Longitudinal neuroimaging studies (e.g., Adolescent Brain Cognitive Development [ABCD] study; https://abcdstudy.org/) are required to unpack the time course of rsFC in people who develop CUD. If indeed such changes did pre-date CUD-onset, patterns of rsFC alterations identified here could contribute to the mapping of biomarkers for CUD, to identify at-risk populations in future research, but could challenge/undermine existing evidence for fMRI differences in CUD relative to control groups.

A strength of this study is its robust methodology, including a focus on a well-defined sample of individuals with moderate-to-severe CUD who have attempted to reduce or cease use. By controlling for confounding factors such as age, sex, and alcohol use, the findings provide a clearer understanding of the specific neural alterations associated with CUD. First, the CUD and control groups did not significantly differ in many sociodemographic and mental health-related variables that can also affect rsFC (e.g., age, anxiety, gender, handedness, and stress; Maron-Katz et al., 2016; Sie et al., 2019; Tomasi & Volkow, 2012, 2024; Xu et al., 2019). Second, confounders that showed significant group differences (i.e., past-30-day alcohol exposure) and variables that exert a major influence on brain function (i.e., age and sex), were adjusted for in all rsFC analyses, minimising their influence on results. Third, participants abstained from cannabis use for at least 12 hours prior to scanning; neither ‘duration of abstinence’ nor ‘THC-COOH: creatinine ng/ml’ correlated with rsFC alterations in the CUD group. Therefore, it was unlikely that acute cannabis effects (i.e., intoxication) confounded results. As well as controlling for potential confounders, another strength of the study was the recruitment of heterogenous participants with a range of depression, anxiety, psychiatric symptoms scores, alcohol, and nicotine use, which is representative of a population with CUD (Onaemo et al., 2021).

Regarding sample size, it has been posited that brain-behaviour correlations are adequately replicable in samples ranging from 20 participants (Spisak et al., 2023) to 42 participants (Makowski et al., 2024). Thus, robust neuroimaging studies with targeted samples with CUD, such as the current experiment, are required to advance the understanding of the neurobiology of CUD. In future, multi-site consortia studies could be utilised to substantially increase power and demonstrate replicability of findings (e.g., ABCD, the ENIGMA Addiction working group; www.enigmaaddictionconsortium.com).

The altered brain networks in CUD identified herein, particularly pallidum-occipital cortex rsFC associated with CUD severity, could be targeted by brain-based interventions for people aiming to gain increased control over, or reduce, their cannabis use. This includes neurofeedback (Russo et al., 2023), Transcranial Magnetic Stimulation (TMS; Kearney-Ramos & Haney, 2021; Mahoney et al., 2020) and mindfulness-based interventions (Lorenzetti et al., 2023), known to reduce alterations of these pathways in SUD. Neurofeedback, which entails the provision of ‘real-time’ feedback during an fMRI scan, regarding brain activation under certain conditions (established previously during substance cravings for alcohol, tobacco, and cocaine; Murphy et al., 2024) allows for the development of personalised neural targets, which the participant can be trained to self-regulate (Martz et al., 2020). rsFC alterations identified herein could be used to guide neural neurofeedback targets. Furthermore, mindfulness-based interventions (Kabat-Zinn, 1991; Kirlic et al., 2021; Witkiewitz et al., 2013) have been linked to alterations of rsFC between brain regions overlapping those implicated in this manuscript, and to a reduction in cigarette smoking (Lorenzetti et al., 2023). Future research could examine if mindfulness interventions targeting core features of CUD mitigate its neural circuitry.

This study is the first fMRI study to examine rsFC in selected ROIs of individuals with moderate-to-severe CUD vs controls, with a robust rationale for the ROIs. We demonstrated greater rsFC across sub-cortical (striatal, basal ganglia) and cortical regions (frontal, occipital, parietal). The changes were partly correlated with the severity of CUD, and they may reflect neuroadaptations in salience pathways that follow the neuroadaptations within the addiction neurocircuitry. Such adaptations in non-intoxicated individuals with CUD may follow documented THC-induced reductions in rsFC and dopamine release within the NAc (Ertl et al., 2023; Wall et al., 2019) – a hypothesis that needs corroboration by experimental studies in CUD before, during, and after THC intoxication.

It was thought that greater putamen/pallidum-occipital/occipito-parietal rsFC, may underlie habitual cannabis use and altered salience processing observed in CUD, in part driven by CUD severity. Furthermore, fronto-striatal rsFC increases in the CUD group may reflect the engagement of salience pathways sensitive to THC exposure, secondary to the effect of THC on dopamine synthesis within the NAc. Longitudinal neuroimaging studies are required to confirm if identified changes predate or follow CUD (or both), and multi-site consortia studies could be instrumental in replicating the findings in individuals with varying CUD severity and in cannabis users without CUD. These findings lend support to prominent neuroscientific theories of addiction insofar as their application to CUD and are likely representative of cannabis-using populations with more severe forms of CUD and comorbid depression and anxiety. Future research could utilise the results herein to develop brain-based interventions for people aiming to change their cannabis use, such as neurofeedback and mindfulness-based intervention.

## Data Availability

The datasets generated during the current study will be available upon reasonable request from Dr Valentina Lorenzetti, Principal Investigator. Data will include relevant group allocations and outcome variables and will be anonymised. Data will be available either as it is published, or on request (following completion of the data collection process, estimated end of 2021). A time limit will not be set on the duration of availability. Data will be shared with anyone who wishes to access it, for meta-analyses or other pre-approved purposes, via email. All participants provided informed consent. All data is de-identified.

## Acknowledgements

We would like to thank all the participants involved in the study, who spent their time supporting our research, and sharing their knowledge and experience. Gratitude is also extended to all past and current staff and student members of the BrainCann Team, who have worked tirelessly on this project since its conception in 2018. This includes Ms Natalie DeBono, Dr Leonie Duehlmeyer, and Dr Penny Hartman for contributing to the management of the setting up of the project. We also acknowledge Ms Stephanie Antonopoulos, Ms Claire Chua, Dr Leonie Duehlmeyer, Dr Alexandra Gaillard, Mr Lachlan Grant, Ms Kirsty Kearney, Dr Magdalena Kowalczyk, Ms Emily Robinson, Ms Elizabeth Sharp, Ms Danielle Tichelaar, and Ms Diny Thomson for their contribution to data collection. We thank Professor Shanlin Fu and the team at the Drugs and Toxicology Group, Centre for Forensic Science, University of Technology Sydney, for conducting urine toxicology analyses.

## Disclosures

Arush Honnedevasthana Arun reports no financial relationships with commercial interests.

Dr Adam Clemente reports no financial relationships with commercial interests.

Emillie Beyer reports no financial relationships with commercial interests.

Dr Lisa-Marie Greenwood reports no financial relationships with commercial interests.

Dr Izelle Labuschagne is the founder and director of Complete Thesis Support, which provides developmental programs for research students.

Professor Valentina Lorenzetti reports no financial relationships with commercial interests.

Dr Victoria Manning was the Founder, CEO, Director and a shareholder of Cognitive Training Solutions Pty Ltd between March 2021 and Aug 2023, which commercialised the SWiPE app which delivers Cognitive Bias Modification to reduce alcohol use.

Dr Eugene McTavish reports no financial relationships with commercial interests.

Dr Govinda Poudel is the founder, director and CTO of BrainCast Pty Ltd which has developed novel brain imaging markers for monitoring brain injury.

Marianna Quinones-Valera reports no financial relationships with commercial interests. Professor Peter Rendell reports no financial relationships with commercial interests.

Dr Hannah Sehl reports no financial relationships with commercial interests.

Dr Chao Suo reports no financial relationships with commercial interests.

Associate Professor Gill Terrett reports no financial relationships with commercial interests.

Dr Hannah Thomson contracts for Syneos Health Learning Solutions, with the Insights and Evidence Generation Team in Patient Insights and Assessment Research (Implementation Science).

## Declaration of funding

Valentina Lorenzetti was supported by an Al and Val Rosenstrauss Research Fellowship (2022-2026), and by a National Health & Medical Research Council (NHMRC) Investigator Grant (2023-2027, ID 2016833) and an Australian Catholic University competitive scheme.

The work within the Neuroscience of Addition and Mental Health Program, Healthy Brain and Mind Research Centre was supported via an ACU competitive scheme.

Hannah Thomson and Hannah Sehl were funded by Australian Government Research Training Program (RTP) Stipend scholarships.

Victoria Manning has received funding from the National Health and Medical Research Council (NHMRC), VicHealth, Department of Health Victoria, the Victorian Responsible Gambling Foundation, the National Centre for Clinical Research on Emerging Drugs (NCCRED), HCF, and philanthropic organisations.

## Previous presentation

Hannah Thomson presented these results via poster presentation on the 25^th^ of July 2022, at the British Association for Psychopharmacology, 2022 London Summer Meeting.

## Authors contribution

- All authors edited the manuscript.
- AHA contributed to fMRI quality checks and analyses, with general direction on the technical aspects from CS and GP and on the theoretical aspects from VL.
- AC managed all the operations of the study.
- EB supported data collection.
- LG provided high-level and ongoing input on all aspects of the study.
- IL supported the setup of the study protocol.
- VL designed and led the study as CI, supervised all students and staff involved, and led all revisions.
- VM provided high-level and ongoing input on the design and conduct of the study and edited the first full draft of the manuscript as well as subsequent drafts.
- EM contributed to statistical analyses of the behavioural data, and provided input on theoretical aspects of the manuscript.
- GP provided high-level and ongoing input on the design and conduct of the study and edited the first full draft of the manuscript as well as subsequent drafts.
- MQ-V supported data collection.
- PR supported the setup of the study protocol.
- HS supported the study setup and data collection.
- CS provided high-level and ongoing input on all aspects of the study with a focus on neuroimaging/statistical analysis and data visualization.
- GT supported the setup of the study protocol.
- HT, under the PhD supervision of VL, CS, and IL, developed the theoretical framework of the manuscript, conducted fMRI quality checks, contributed to the neuroimaging data analysis, drove statistical behavioural analysis, created the first draft and integrated subsequent revisions.

## Footnotes

a One participant who turned 56 between screening and testing was retained in the sample.

## Notes

### Clinical Trial

ISRCTN ID: 76056942

