## Supplemental Information for "The addiction neurocircuitry during Resting-State Functional Connectivity fMRI in people with Cannabis User Disorder who tried to cut down or quit"

### SUPPLEMENTARY INFORMATION

#### TABLE OF CONTENTS

1. INCLUSION AND EXCLUSION CRITERIA - 2 -
2. PROCEDURE - 3 -
3. fMRI ACQUISITION AND PROCESSING - 4 -
4. SUPPLEMENTARY FIGURE 1. **Monash Biomedical Imaging Magnetic Resonance Imaging (MRI) Screening and Information Form** - 7 -
5. REFERENCES - 8 -
6. SUPPLEMENTARY TABLE 1. **CUD group correlations between rsFC pairs showing group differences (rsFC beta values) correlated with metrics of cannabis exposure and related problems, depression scores, and nicotine dependency, using Bonferroni adjusted alpha threshold of p<.001 (.05 ÷ 45)** - 10 -

1. INCLUSION AND EXCLUSION CRITERIA

Unless otherwise specified, inclusion and exclusion criteria were confirmed by participant self-report and the use of a comprehensive online screening survey followed by detailed and focused screening over the phone.

Inclusion Criteria

Inclusion criteria for all participants were: (1) age between 18 years and 55 years; (2) having normal-to-corrected vision; (3) fluency in English; and (4) ability to attend testing sessions in person.

For people with CUD only: (1) use of cannabis on a daily or almost daily basis for ≥ 12 months; (2) an attempt to quit or to reduce cannabis use at least once within the past 24 months; and (3) a diagnosis of moderate-to-severe CUD, confirmed by the endorsement of ≥ 4 CUD symptoms from the DSM-5 (American Psychiatric Association [APA], 2013), measured using the Structured Clinical Interview of DSM-5 – research version (SCID-5-RV; First et al., 2015).

Exclusion Criteria

Exclusion criteria for all participants were: (1) MRI contraindications, measured using the Monash Biomedical Imaging Magnetic Resonance Imaging (MRI) Screening and Information Form (*Supplementary Figure 1*); (2) unwillingness to refrain from any illicit substance and/or alcohol use in the 12 hours before testing (confirmed by self-report upon arrival at session); (3) current use of prescription medication that affects the central nervous system, except anti-depressants and anxiolytics due to prevalence of elevated depression/anxiety levels in CUD; (4) current suicidal ideation or history of any diagnosed psychiatric disorders, with the exception for depression and anxiety disorders due to the high comorbidity with CUD, as confirmed using The MINI International Neuropsychiatric Interview 7.0.2 (MINI; Lecrubier et al., 1998; Lecrubier et al., 1997; Sheehan et al., 2015; Sheehan et al., 1998); (5) history of any neurological disorders or major medical conditions (e.g., epilepsy, stroke, migraine, etc.); (6) history of acquired or traumatic brain injury or loss of consciousness > 5 minutes; (7) full scale intelligence quotient (FSIQ) estimate score < 70, confirmed using the Wechsler Abbreviated Scale of Intelligence – second edition (WASI-II; Wechsler, 2011); (8) current pregnancy and/or breastfeeding; (9) history of significant and regular mindfulness practice, defined as regular engagement in mindfulness practices over any extended period of time; or (10) significant dependence on alcohol, confirmed by a score > 19 on the Alcohol Use Disorders Identification Test (AUDIT; Babor et al., 2018).

For people with CUD only, exclusion criteria were: (1) significant dependence on or exposure to substances other than cannabis or tobacco, threshold at >50 occasions of use over a 2-year period in the past 10 years; or (2) or use of any illicit drug other than cannabis in the 4 weeks prior to testing.

For controls only, exclusion criteria were: (1) significant dependence on or exposure to substances other than tobacco, threshold at >50 occasions of use over a 2-year period in the past 10 years; (2) use of any illicit drug in the 4 weeks prior to testing; (3) use of cannabis at any stage in the 12 months prior to testing; or (4) > 50 lifetime uses of cannabis.

2. PROCEDURE

Members of the community who were interested in the study were directed to an online screening survey to determine their eligibility against inclusion and exclusion criteria (~30-minutes, 9,045 respondents). This was followed by a phone screener to confirm eligibility of potentially suitable participants (lasting ~10 minutes to ~1 hour; ~450 prospective people with CUD and ~800 prospective controls were called). A detailed list of the tools used for the online and telephone screening is outlined in the study pre-registration (ISRCTN ID: 76056942). Eligible participants then attended face-to-face data collection at Monash Biomedical Imaging facility, Clayton, Victoria, Australia.

The testing session lasted approximately 4-6 hours (including the collection of measures beyond the scope of this study). Participants completed an assessment battery to profile socio-demographic data, substance use, mental health, and cognition, comprising questionnaires, face-to-face semi-structured interviews, standardised cognitive testing, as well as MRI scanning and provision of urine samples. All questionnaires used for participants' screening and face-to-face testing were administered via Qualtrics Software, version 2019-2022 (Qualtrics, Provo, UT).

This study was part of a larger longitudinal study. At the end of testing, participants were offered the opportunity to debrief and receive reimbursement in the form of a Coles Myer voucher to the value of AUD $100 for controls, and of AUD $150 for people with CUD, as they underwent additional testing to address research questions beyond the scope of this study. All participants were also offered a picture from a single frame from their T1-weighted (T1w) scan.

3. fMRI ACQUISITION AND PROCESSING

**MRI Task Setup**

Prior to entering the scanner participants were requested to stay awake; the tester checked in real-time that participants kept their eyes open throughout the scan, via an MRI-compatible camera placed inside the MRI scanner.

**rsFC fMRI Task Instructions**

Inside the MRI scanner, and prior to the resting-state scan acquisition, participants were instructed verbatim by the researcher and via written instructions: “The next scan will take about 10 minutes. Keep your eyes open, try not to think about anything in particular. Stay relaxed and try to keep your head still”. Through the resting-state scan, participants were shown a fixation cross (white cross on black background) via a mirror placed inside the MRI scanner.

**MRI Data Acquisition Parameters**

Participants were scanned using the same group of experienced radiographers at the Monash Biomedical Imaging facility in Clayton, Victoria. All participants were scanned on the same Siemens Skyra 3 Tesla MRI scanner using a 32-channel head coil. *T1-weighted (T1w)* scans were acquired using magnetization-prepared rapid gradient-echo (MP-RAGE) sequence, with the following acquisition parameters: TE = 2.07ms, TR = 2300ms, flip angle = 9°, 192 sagittal slices without gap, field of view 256 x 256mm, yielding a 1 x 1 x 1mm resolution, for a total acquisition time of 5 minutes. *Resting-state* fMRI scans (189 volumes) using T2* weighted Echo Planar Imaging (EPI) were acquired using the following parameters: TE = 30ms, TR = 2500ms, flip angle = 90°, 44 slices without gap, field of view 192 x 192mm, matrix = 64, voxel size 3mm3, and a total acquisition time of 8 minutes. Slice based acceleration Generalised Autocalibrating Partially Parallel Acquisitions (GRAPPA) = 2 was applied. The Siemens system includes an additional two ‘dummy scans’ at the beginning of the acquisition to allow the magnetization to stabilise to a steady state. These two scans were not collected for any data analysis.

**MRI Data Handling**

All MRI data were directly exported from the scanner to Monash Biomedical Imaging-XNAT (XNAT website, private server), where it was stored and backed up in Digital Imaging and Communications in Medicine (DICOM) format. Raw format (i.e., DICOM) data were downloaded from the XNAT server and converted into Brain Imaging Data Structure (BIDS) format using dcm2niix (v1.0.20201102) for further analysis. All imaging data processing and analysis were performed on a cloud-based cluster-computational platform, MASSIVE (massive.org.au; Goscinski et al., 2014). The pre- and post-processing was conducted using CONN toolbox 20.b (www.nitrc.org/projects/conn, RRID:SCR_009550; Whitfield-Gabrieli & Nieto-Castanon, 2012), based on SPM12 on Matlab (2018a.r7487), which was pre-installed on MASSIVE.

**MRI Data Pre-Processing**

All validated data were imported in BIDS format, then underwent a standard pre-processing using CONN toolbox 20.b, including: (1) slice timing with interleaved slice order; (2) realignment and generation of motion parameters; (3) ARtifact-detection Tools (ART)-based outlier detection with intermediate settings (default 97$^{th}$ percentile in normative sample); (4) co-registered fMRI data with T1w images; (5) segmentations of T1w images; (6) normalising T1w images to Montreal Neurological Institute (MNI) space (standard space) and normalising fMRI to MNI space with the same parameters; (7) default nuisance factors removal including six realignment parameters, six first order temporal derivatives, signal from white matter, cerebral spinal fluid, and whole brain, (8) smoothing with 6mm kernel; and (9) temporal bandpass filtering (0.008-0.09). Default CONN toolbox 20.b pre-processing steps also include a combination of aCompCor, scrubbing, and motion correction. fMRI was then resampled to 2 x 2 x 2mm isotropic voxels. Quality Assurance reports were generated and manually reviewed by authors HT and CS. Stringent criteria for detecting motion outliers, as outlined by Parkes et al. (2018), was followed, whereby limits of >0.25mm mean framewise displacement (mFD) and >5mm maximum framewise displacement were set; no participants violated these criteria.

**MRI Data Post-Processing**

The seed-based rsFC maps for the significant three seeds were used for further post-processing. The location of the seeds was determined by the default Harvard-Oxford atlas (132 cortical and subcortical regions; Desikan et al., 2006). Seed-based functional connectivity maps were generated using CONN toolbox 20.b. The group effect of the seed-based connectivity map was analysed using a General Linear Model (GLM) model using group as a factor (between subjects, two-group test). Covariates were age, sex, and total number of alcohol standard drinks over the past 30 days. Voxel wise multiple comparison error correction was applied, specifically a cluster-level False Discovery Rate (FDR) correction, p-corrected $< 0.05$ was applied, with default initial $p < 0.001$. To make the results more robust, a Benjamini-Hochberg procedure was additionally applied to alpha values to decrease the false discovery rate following the examination of multiple ROIs (Benjamini & Hochberg, 1995).

4. SUPPLEMENTARY FIGURE 1. ***Monash Biomedical Imaging Magnetic Resonance Imaging (MRI) Screening and Information Form***

Have you ever had any eye injury caused by metal? ....................................................................................NO / YES
If YES:
Did you see a doctor at the time? ....................................................................................NO / YES
Did they remove the foreign body? ....................................................................................NO / YES
Did they tell you that they got it all out? ....................................................................................NO / YES
Was this the last injury involving metal? ....................................................................................NO / YES
Are you pregnant, suspect you may be pregnant or breastfeeding?....................................................NO / YES

**Do You Have (Or Have You Ever Had):**
A Cardiac Pacemaker/stent/defibrillator/wire.....................................................NO / YES
Any heart operation or valve replacement.....................................................NO / YES
Any Brain operation .....................................................NO / YES
Abdominal Aneurysm repair or IVC filter.....................................................NO / YES
Brain Aneurysm Clips.....................................................NO / YES
Deep Brain Stimulator.....................................................NO / YES
Brain Shunt Tube.....................................................NO / YES
If YES, is it programmable .....................................................NO / YES
Any Ear operations /cochlear or stapes implants.....................................................NO / YES
Implanted drug infusion devices.....................................................NO / YES
Neuro or Bone growth stimulator.....................................................NO / YES
Shrapnel, bullet, gunshot.....................................................NO / YES
Any stents, vascular, oesophageal or biliary.....................................................NO / YES
Any Surgical clips/wire sutures/screws/mesh/prosthesis. .....................................................NO / YES
Joint Replacement or Prosthesis.....................................................NO / YES

**Do You Have:**
Ocular prosthesis (eye implants).....................................................NO / YES
A Swan-Ganz Catheter .....................................................NO / YES
Skin patches .....................................................NO / YES
Intrauterine device (IUD).....................................................NO / YES
A penile prosthesis .....................................................NO / YES
Any other implant, or breast tissue expander .....................................................NO / YES
Tattooed eyelids or tattoos.....................................................NO / YES
Hearing Aid .....................................................NO / YES
Removable dentures.....................................................NO / YES
Any Piercings or braces that CANNOT be removed.....................................................NO/ YES
Hair Extensions .....................................................NO / YES

**Have You:** **What? / When?**
Had an operation or procedure within the last 8 weeks ..............................NO / YES ................................
Had a history of seizures or epilepsy .....................................................NO / YES

6. SUPPLEMENTARY TABLE 1.

**CUD group correlations between rsFC pairs showing group differences (rsFC beta values) correlated with metrics of cannabis exposure and related problems, depression scores, and nicotine dependency, using Bonferroni adjusted alpha threshold of p<.001 (.05 ÷ 45)**

| Seed region | Cluster region/s | | SCID-5-RV | CUDIT-R | Grams, past 30 days | THC-COOH: creatinine ng/ml | Hours since last use | Age of reg. onset | Duration of reg. use | BDI-II[a] | FTND |
|---|---|---|---|---|---|---|---|---|---|---|---|
| *right* NAc | frontal | $r_s$ | -.10 | .03 | -.02 | -.03 | -.17 | .08 | .14 | .02 | -.13 |
| | | p | .45 | .83 | .89 | .83 | .19 | .54 | .28 | .88 | .30 |
| *left* putamen | occipito-parietal | $r_s$ | .10 | -.10 | .04 | -.11 | .25 | -.28 | .08 | -.04 | -.11 |
| | | p | .45 | .46 | .73 | .38 | .05 | .02 | .53 | .78 | .41 |
| *left* pallidum | occipital | $r_s$ | -.16 | -.42 | .12 | .11 | .13 | .01 | .02 | -.43 | -.10 |
| | | p | .20 | **<.001***** | .34 | .38 | .31 | .93 | .86 | **<.001***** | .44 |
| *right* pallidum | occipital | $r_s$ | -.02 | .10 | -.02 | .10 | .18 | .10 | -.18 | -.08 | -.15 |
| | | p | .87 | .42 | .90 | .98 | .15 | .42 | .15 | .55 | .24 |
| | occipital | $r_s$ | .12 | -.05 | .08 | .04 | -.03 | .10 | -.14 | .05 | -.06 |
| | | p | .35 | .69 | .54 | .74 | .83 | .44 | .26 | .68 | .65 |

BDI-II, Beck Depression Inventory – second edition; CUDIT-R, Cannabis Use Disorder Identification Test-Revised; FTND, Fagerström Test for Nicotine Dependence; NAc, nucleus accumbens; $r_s$, Spearman's Rho; SCID-5-RV, Structured Clinical Interview of DSM-5 – research version; THC-COOH: creatinine ng/ml, 11-nor-9-carboxy-$\Delta^9$-tetrahydrocannabinol: creatinine per millilitre of urine

*Note*: Bolded p-values represent statistically significant, moderate ($r_s$: 0.40 – 0.69) correlations *****<.001**

[a] Due to collinearity with the BDI-II (Beck et al., 1996), CAPE (Stefanis et al., 2002) subscale scores were not included.
